# Where Do I Belong? Searching for fit in an unseen specialty: medical students’ paths to Youth Health Care

**DOI:** 10.64898/2026.07.14.26358057

**Authors:** Janneke Z. Muyselaar-Jellema, Karen D. Könings, Anne van Eijk, Jessica C. Kiefte-de Jong, Vera Nierkens

## Abstract

**Introduction:** As healthcare systems increasingly shift toward prevention and community-based care, the demand for physicians in extramural specialties continues to grow. Yet, a mismatch persists between workforce needs and medical students’ career aspirations. Little is known about how medical students and trainees develop an interest in extramural specialties such as youth health care (YHC). This study explores trainees’ trajectories toward becoming a youth health care physician (YHCP).

**Methods:** We conducted a qualitative study using semi-structured online interviews with fourteen YHCPs in training. We combined an inductive and deductive approach, applying the person-environment (PE) fit framework to explore participants’ evolving experiences of fit and misfit.

**Results:** Participants described growing misfit with clinical culture of medical school (i.e. the ‘hidden curriculum’), particularly during hospital-based clerkships, combined with limited exposure to YHC. For some, this misfit extended to doubts about becoming a doctor. Over time, participants developed a sense of fit and belonging within YHC, either directly or after exploring other specialties including extramural specialties.

**Discussion:** These findings reframe specialty choice as a longitudinal search for belonging and alignment, in which trainees iteratively explore, evaluate, and refine their sense of fit across contexts. Clerkships serve as key sites for testing fit, yet also expose learners to the clinical culture, including the hidden curriculum. Broadening exposure and supporting reflective fit processes may encourage more medical students to choose extramural specialties, ultimately fostering a more balanced and sustainable alignment of the medical workforce.

## Introduction

There is a growing global need for physicians in extramural specialties to meet increasing demands for community-based care and the shift towards prevention [1]. Ageing populations require more holistic and integrated care, while reducing the demand for highly specialised hospital care. A persistent mismatch exists between medical students’ career aspirations and workforce needs [2]. However, the processes underlying students’ interest in extramural specialties, such as youth health care YHC, remain poorly understood.

Career intentions develop dynamically during medical training, particularly during clinical clerkships, a period marked by intensive career exploration and decision-making stress [3]. Specialty preferences evolve over time [4-6] with the most pronounced changes occurring during clinical exposure [7, 8]. Existing literature conceptualises specialty choice as the result of an interplay between personal values, career needs, and perceptions of specialty characteristics [9-12], with experiential factors such as role models and perceived work–life balance further shaping preferences [13-16]. These processes can be understood through a PE fit perspective, which emphasises alignment between individuals and their work environment [17]. Within this model as shown in Figure 1, multiple dimensions of fit — person–job (PJ), person–organisation (PO), person–supervisor (PS), and person–group (PG) — have been described [18]. PJ characteristics relate to features of the job itself, such as the patient population, the type of work, and working hours. PG characteristics concern colleagues and team dynamics, whereas PO characteristics encompass organizational values, size, and hierarchical structure. While perceived fit is associated with job satisfaction and retention [19, 20], misfit reflects a deeper sense of incompatibility between individual and environment [21].

**Figure 1:**
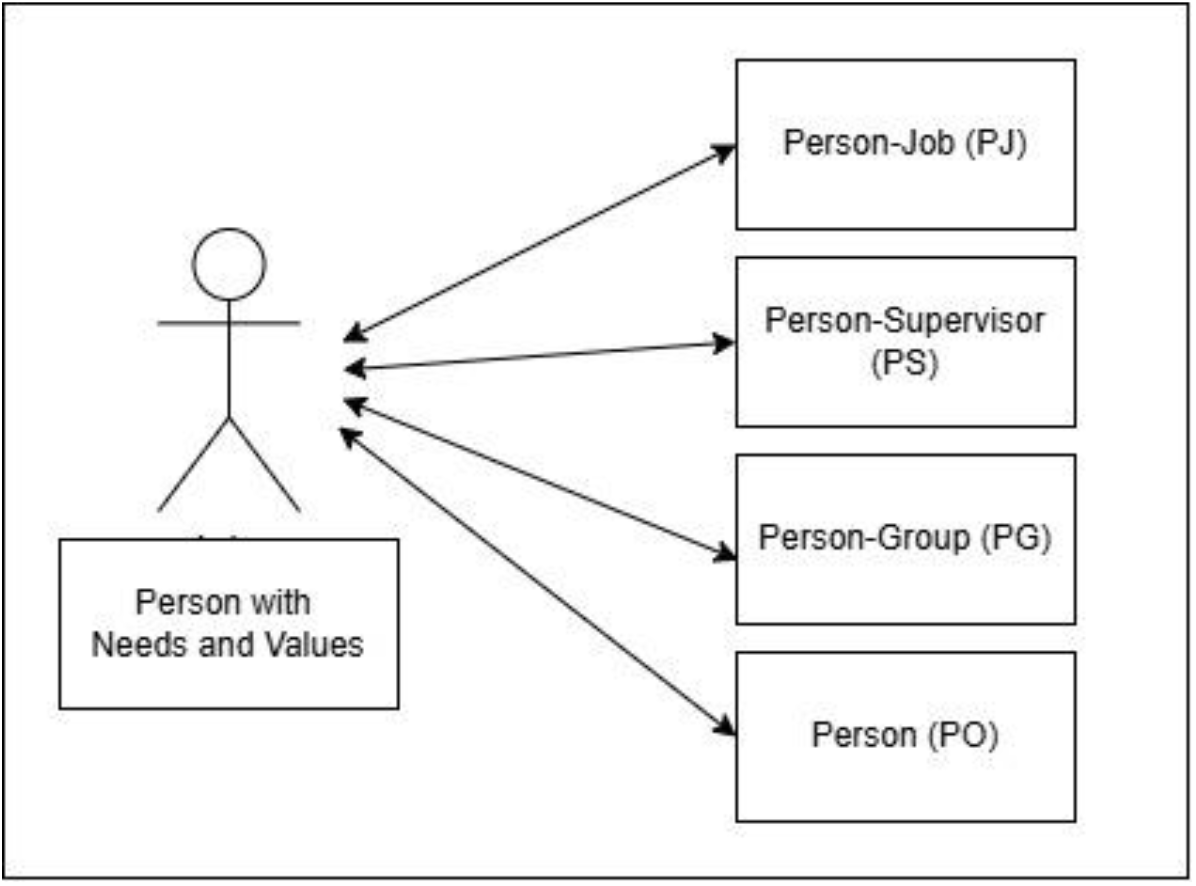
Person-Environment (PE) fit model: showing the four different dimensions of fit.

There is limited knowledge of how experiences of fit and misfit shape the development of interest in extramural specialties, and how these experiences interact with emerging career intentions in community-based settings. Better understanding students’ career decision-making can contribute to better recruitment into community-based specialties. This study aims to explore how individuals experience and navigate their trajectories toward becoming a YHC. The research question is: How do individuals experience and navigate their trajectories toward becoming a YHCP, including moments of fit and misfit?

## Methods

We conducted a qualitative study based on a constructivist research paradigm. In this philosophical framework, reality can be understood through people’s experiences and interpretations.

### Participants

Our study population comprised of youth health care physician (YHCP) trainees in the Netherlands. In this context, we define YHCP, as the extramural specialty focusing on preventive healthcare for the ages 0-18 years old as provided within the Dutch Youth Health Care (Jeugdgezondheidszorg, JGZ) [22]. In 2023, a total of 236 YHCP trainees were enrolled in the two-year postgraduate training programme.

### Materials

The semi-structured interview focused on the process of career intention and career choice. The interview guide was informed by the conceptual framework developed by Pfarrwaller et al. [23]. We asked participants how medical school experiences shaped their specialty intention. Were there specific people or situations that influenced your choice? The interview guide (Appendix 1) was pilot tested by AvE and small adjustments were made concerning the question order.

### Procedure

Recruitment took place nationwide via YHCP training institutions, a Facebook group, and snowball sampling. Participation was voluntary and without incentives. Seventeen individuals responded (16 female, 1 male); four did not participate for practical reasons. To improve gender balance, one additional male was purposively recruited. All participants provided written informed consent prior to the interviews. Data collection and analysis were iterative. Fourteen participants were interviewed online via Microsoft Teams (March–July 2024) by AvE and JM. Interviews lasted 36–52 minutes, were audio-recorded, and transcribed using Amberscript [24].

### Data analysis

Thematic analysis was used to code and organise the data into themes until thematic sufficiency was reached. Two researchers (AvE, JM) coded the transcripts and resolved differences through discussion. Analysis combined theory-informed inductive [25] and deductive approaches, guided by the PE fit model [17]. Selected quotations were translated into English; numbers indicate unique participant codes. The study was approved by the N-Wmo Ethical Research Board of the PHEG department, LUMC (23-3099), the Netherlands.

### Reflexivity

Our research team includes a YHCP, medical teacher and PhD candidate in medical education (JM), a YHCP trainee (AvE), a health scientist, medical teacher and qualitative researcher (VN), associate professor and psychologist (KK) and professor of population health (JK), all working at Dutch medical universities. These various perspectives were negotiated in the team’s discussions regarding data interpretation and meaning making, inherent to constructivist approaches.

## Results

Fourteen participants were interviewed (12 female, 2 male), with a mean age of 33.9 years (range 26– 44). Nearly all completed medical school at different universities in the Netherlands; one was a refugee physician with prior experience as a pediatrician in multiple countries. About half had worked in various specialties (including surgery, pediatrics, rehabilitation medicine, elderly care, and clinical genetics), while the remainder entered YHC directly after graduation. Participants’ accounts reflected a processual trajectory from perceived misfit to a sense of belonging, characterised by four themes: exposure to clinical culture in medical school, increasing misfit during hospital-based clerkships, limited exposure to non-hospital career options such as YHC, and the gradual emergence of fit within YHC.

### Experiencing the Clinical Culture of Medical School

Clinical culture of medical school shaped participants’ early career intentions. They experienced a normative environment in which pursuing a hospital-based specialty was implicitly expected. As a participant stated: “You are going to study medicine, so you will become a hospital specialist.” (R9) This expectation was conveyed through both formal and informal elements of training, including role modelling by senior physicians, a curricular focus on hospital medicine, and everyday interactions with supervisors and peers. As a result, participants’ initial conception of a physician was largely limited to the traditional, hospital-based role, often symbolized by the white coat. Accordingly, many participants initially aspired to become pediatricians, emergency physicians, or psychiatrists, while others were drawn to fields such as hematology, genetics, or immunology, often because they had enjoyed the teaching and lectures in these areas. Several participants described feeling pressure to align their aspirations with this dominant clinical culture. Participants encountered clinical culture in two main contexts: the pre-clinical phase (lectures and classroom-based learning) and clerkships (work-based learning across specialties within and beyond hospital settings). Across both contexts, they reported a sense of misfit. The clinical culture was absorbed by the students: “The more you specialize, the “cooler” it seems, and I really went along with that whole rush”. (R14) Participants described the culture as strongly performance-oriented, with certain highly specialized careers positioned as the ultimate markers of success, as illustrated by one participant: “In medical school there’s basically this really old performance-driven culture. Right from the first lectures, you’ll have a cardiothoracic surgeon standing there, in a nice white coat, telling their story, and everyone’s like: oh wow, this is cool. This is what I want to be! That kind of gets framed as the ultimate goal. And that culture basically never really disappears” (R14).

For some participants, the dominant clinical culture reinforced uncertainty and self-doubt about their career preferences, while others actively resisted it by seeking experiences outside the hospital. Participants who considered non-hospital careers often felt misunderstood and isolated, particularly when surrounded by peers with clearly defined ambitions to become hospital specialists. One participant recalled being viewed as a “loser” in a mentoring group because she did not aspire to a clinical specialty (R13). Participants also described having to justify their career intentions to peers, faculty, and family, which sometimes delayed career decisions. As one participant explained, common negative perceptions and limited understanding of the field made it feel as though she was “always having to justify” her choice (R4).

Several participants perceived alternative career paths such as YHC as less visible and less prestigious within medical education. They also described derogatory comments by faculty and specialists, such as pediatricians referring to YHCPs as “doll doctors” (R9). Alongside uncertainty, participants reported limited support and guidance in career decision-making. They attributed this to the predominantly hospital-based backgrounds of faculty, who were perceived as having limited knowledge of careers beyond the clinical setting. As a result, participants felt they had to “figure it out on their own”, with little attention to “personal development and career choice” (R7). One participant described a mismatch with a mentor whose highly hospital-oriented career trajectory left little room to discuss doubts about becoming a doctor: “During the clinical rotations, my mentor was an ICU doctor who worked fulltime, had four kids, was doing a PhD at an academic center, and did a million other things, and absolutely loved it. So that wasn’t really someone I could… [turn to]” (R13).

### Experiencing Misfit in the Hospital Environment

Upon transitioning into clerkships and encountering the realities of hospital life and culture, some participants started to question whether they still wanted to become a doctor at all. “I actually started seriously doubting whether I even wanted to be a doctor at all. None of the hospital rotations I’d done up to that point were particularly interesting, and I realized I didn’t really enjoy working in the hospital either. So I really started doubting, like… okay, if not this, then what do I want?” (R1) One participant did not initially experience a misfit in the hospital and thus represented an exception. However, despite enjoying hospital work after graduation, prolonged working hours and an unfavorable work–life balance led this participant to explore a career outside the hospital setting. “I just didn’t want that kind of life. I had way too much FOMO [fear of missing out]. Whenever everyone else was doing fun things, I’d be stuck working a night shift” (R8).

Within the PE fit model, alignment between individuals’ needs and values and those of their work environment is central. Participants reported a lack of such alignment during hospital-based clerkships, describing a sense of misfit and an inability to “be themselves.” For some, this misfit was immediate during their first clerkships, while others experienced it later or after graduation. This perceived misfit heightened participants’ awareness of their own needs, preferences, and values, shaping what they sought in a future career path: “I really go with my gut, and I felt it immediately: this — [the hospital] — just isn’t my place. I know when I feel comfortable somewhere and what it’s like when I’m really able to thrive. And I didn’t feel like myself there at all, and I also felt like my strengths just didn’t really come through there” (R5).

Participants experienced misfit across all four dimensions of the PE fit model (PJ, PS, PG, and PO), although misfit with the organisation (PO) was most prominent. As one participant explained: “I think it’s a combination of things — the atmosphere in the hospital, the hierarchy, the scale of it all, the workload, the irregular shifts. Yes, all things that just don’t really suit me” (R7). Additional examples included a lack of interest in caring for sick patients or children, emergency care, on-call duties, and hospital work more broadly (PJ); mismatches with supervisors and their expectations (PS); and experiences of professional rivalry, hierarchy, and an impersonal work environment (PG). Together, these experiences led participants to rule out hospital-based specialties as career options.

### Exploring Options and Limited Awareness

After deciding not to work in a hospital, participants described a period of exploration characterized by uncertainty. Awareness of alternative career pathways within medicine was limited, and General Practice was one of the few non-hospital specialties known to them. While confident about what they did not want, they remained unsure about which specialty to pursue. This phase—lasting from months to several years—involved repeated trial and error and was perceived as costly in time and energy: “It was very much a search. For over seven years I tried all sorts of things, ……. you do learn something everywhere you go” (R7). Experiences of misfit prompted an active search for better alignment with their personal needs and values as well as with dimensions of PG and PO. Participants described a gradual process of reflective exploration, as one participant illustrates: “I was struggling with these questions: what do I want to do? And what actually suits me?” (R4)

Participants reported limited awareness of YHC during medical school, where the specialty was not explicitly embedded in the curriculum and remained largely invisible as a career option. Exposure was uncommon and typically restricted to brief, non-mandatory clerkships, which were perceived as low priority. “I did do a clerkship in YHC, but I cannot remember it very well. It did not make much of an impression on me at the time. Two weeks, I think It was the clerkship that was generally considered the least important and the least enjoyable.” (R10) As a result, many participants encountered the specialty only incidentally—through informal contacts, personal healthcare experiences, or media (e.g., podcasts and online videos)—or only after graduation. Some described this late discovery as unexpected or even revelatory (R1). Overall, opportunities to actively consider YHC were scarce, contributing to a delayed understanding of its scope in career decision-making.

### Finding fit, belonging and connection in YHC

Even though several participants had limited awareness of YHC, some participants chose YHC immediately after graduation (pathway 1; Figure 2). These participants described positive experiences with the specialty during medical school, which helped them recognize an early sense of fit and enter postgraduate training. In contrast, participants who did not recall prior exposure to YHC (pathway 2) described the specialty as largely unknown to them. As a result, they often first explored other specialties before ultimately identifying YHC as the best fit.

**Figure 2:**
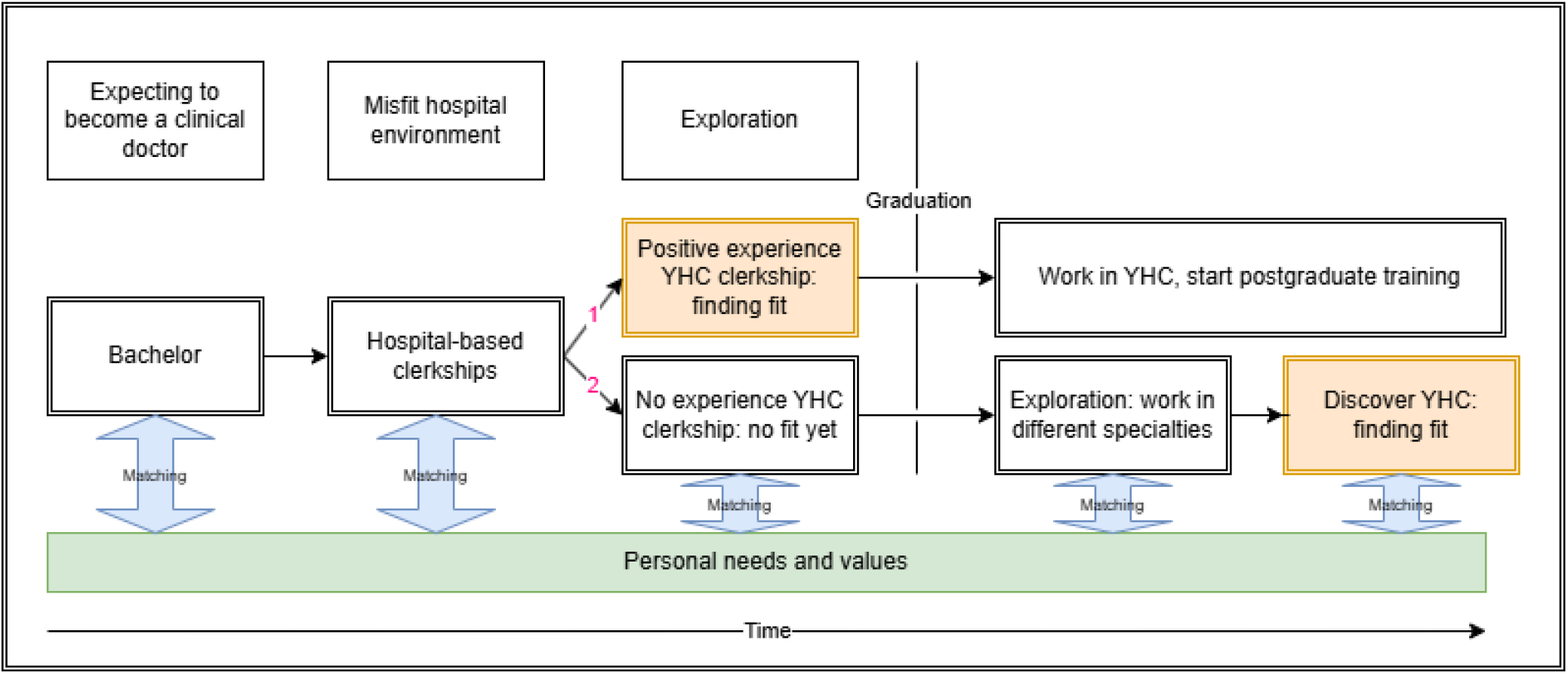
Visualisation of the two trajectories and the timing of finding fit. Note. Pathway 1: Experience YHC clerkship; Pathway 2: no experience of YHC clerkship

In YHC, participants experienced a strong alignment between their personal values and needs, including the desire for family life, financial security, and working close to home. They described a strong sense of fit across the PJ, PG, and PO dimensions, valuing regular working hours, smaller teams, supportive collegial relationships, low hierarchical structures, and a positive work atmosphere. As one participant explained: “I much prefer a smaller team where I know everyone, regular working hours, a nice atmosphere without much hierarchy—or preferably none at all—where people are friendly, learn from one another, where it feels relaxed and enjoyable, and where the workload [is lower]” (R7).

Participants were also attracted to key job characteristics of YHC, such as working with children, focusing on prevention, adopting a contextual approach to care, and engaging in diverse activities. This alignment was commonly described as a sense of belonging. As one participant noted: “In a very enthusiastic team that shared a lot of my ambitions, shared interests… I really felt at home; these are my kind of people” (R13). Participants reported feeling able to be themselves, enjoying their work, thriving professionally, and achieving a satisfactory work–life balance. Their sense of fit was further affirmed by others’ observations that they seemed to have found the right place professionally, as well as by their own growing enthusiasm for the specialty. As one participant reflected: “I could really feel it—and others noticed it too—that I was really thriving there and getting genuinely excited about it. It just really felt like I belonged there” (R11). Overall, participants experienced YHC as a setting in which they could fully express themselves and engage meaningfully in their work. As one participant concluded, “out of everything I’d tried, YHC was actually the best fit for me” (R7).

## Discussion

This study explored how career intentions develop over time among trainees pursuing YHC. Participants’ accounts portray specialty choice not as a linear progression toward a predefined career path, but as a prolonged and often solitary process of exploration, characterized by recurring experiences of fit and misfit across all four dimensions of the PE fit model (PJ, PS, PG, and PO). Over time, these experiences enabled trainees to clarify their preferences and ultimately develop a sense of belonging within YHC.

Consistent with Thomas et al. [26], our findings demonstrate the value of the PE fit model for understanding career preference development. At the same time, our findings suggest that fit is not a static state but a dynamic process, shaped by ongoing reflection and experiences across different training contexts. Clerkships, in particular, provide opportunities for students to explore and evaluate their fit within different clinical settings.

Our findings align with wider literature framing career intention development as a process of testing for fit, in which individuals actively evaluate alignment between personal values and professional environments [26, 27]. Importantly, this testing phase appears to precede a sense of belonging, as also described by Pfarrwaller [28] among postgraduate trainees with an interest in primary care and by Singh et al. [29] among medical students who revised their career intentions. From a PE fit perspective, this sense of belonging reflects PG fit, arising from alignment with the norms, values, and identities of the community working in YHC. In our findings, such group-level fit did not merely signal satisfaction or stability, but appeared to create conditions for thriving, as participants described renewed professional energy and growth once they felt recognized as legitimate members of the community.

Notably, this study identified a form of misfit that extends beyond specialty choice. For some trainees, clerkship experiences prompted doubts not only about which specialty to pursue, but about whether they belonged in medicine at all. These doubts appeared to stem from a mismatch with dominant hospital-based images of what it means to be a doctor. The white coat, long associated with hospital-based medicine, shapes medical students’ evaluations of physicians [30], yet its role in shaping students’ own professional aspirations remains underestimated. Our findings suggest that participants questioned whether they belonged within dominant conceptions of being a doctor, often embodied in traditional white coat–centred clinical cultures. Fit was restored when participants encountered alternative professional identities in extramural specialties such as YHC that better aligned with their values, preferences, and ways of relating to children and parents. The PJ fit aligns with previous research, in which YHCPs identified working with young patients, preventive care, and communication as the most attractive aspects of the profession [22]. Rather than serving as a fallback option, YHC provided a meaningful professional identity through which participants could remain in medicine, underscoring the importance of recognising multiple legitimate ways of being a doctor.

Our findings illustrate how exposure to the dominant clinical culture of medical school shapes early career intentions. Recent Dutch qualitative research similarly identified both medical school and hospital culture as influential in medical students’ career choices [31]. As articulated in Pfarrwaller’s conceptual model [23] and supported by Thomas et al. [26], medical school culture exerts an influence on specialty choice through both formal and informal learning experiences. These influences are closely linked to the hidden curriculum, which has been described as an unavoidable and powerful force shaping how medical trainees develop their professional identities [32]. The hidden curriculum refers to the implicit transmission of values, norms, and behavioural expectations that occurs alongside the formal curriculum, but remains largely unacknowledged and unarticulated within official educational structures [33]. Consistent with literature on specialty disrespect as part of the hidden curriculum (26,27), participants’ reports of “bad mouthing” show how implicit hierarchies shape specialty perceptions and may divert trainees from particular career paths.

Ideally, medical schools can play an active role in mitigating the potentially harmful effects of the hidden curriculum by promoting reflective practice and explicitly naming and discussing these implicit cultural messages with faculty and students. Clerkships provide opportunities for students to explore and evaluate their fit within different clinical settings. By encouraging structured reflection on how students’ personal needs, values, interests, and preferences align with the clinical demands, cultures, and working conditions of different specialties, clerkships can promote more informed and sustainable career decision-making. Previous work suggests that structured opportunities for reflection and open dialogue enable learners to recognize, critically appraise, and contextualize hidden curricular influences, thereby reducing their negative impact on professional identity formation and career decision-making [34-36]. Encouraging faculty to communicate positively about the full breadth of available specialty options - including extramural and community-based practice - and to actively support students in testing for fit, may further help counteract implicit hierarchical messaging and broaden students’ perceptions of viable medical careers [37]. Such exposure may be especially important for students who experience a misfit with the hospital environment, as awareness of alternative professional identities can support continued engagement with medicine and facilitate career choices that better align with their values and strengths. This is increasingly relevant given the growing societal need for a strong community-based medical workforce.

A theoretical implication of this study is that career preference development may be more strongly influenced by clinical culture than current theories explicitly acknowledge. Our findings suggest that the clinical culture of medical school—including its hierarchies, role models, and implicit messages about specialty prestige—shapes how trainees evaluate specialties and their own sense of fit within medicine. While socio-ecological models such as Pfarrwaller et al. [23] recognize the multiple contextual levels surrounding learners, our findings highlight how clinical culture operates across these levels, influencing which career paths are visible, valued, and perceived as legitimate. Future models may therefore benefit from more explicitly incorporating cultural influences and system-level factors on career intention development.

## Methodological considerations

To our knowledge, this is the first study to apply the PE fit model to examine trajectories toward a career in YHC, while also providing in-depth insight into participants’ experiences along these pathways. Additionally, our sample included participants from across different regions of the Netherlands and from multiple universities, enhancing the diversity and potential generalizability of our findings.

Several limitations should be considered when interpreting these findings. Our findings are situated within the Dutch medical education context, where graduates may transition directly into postgraduate training or first gain work experience within a specialty, which influences how career choices and fit are experienced and articulated. Another potential limitation is recall bias, as participants were asked to reflect retrospectively on their career trajectories. These reflections may slightly differ from experiences and perceptions at the time choices were made, which could have influenced the nuance of the data. Regarding the participants, although we actively sought male participants, only two were included; one had a markedly different trajectory as a refugee who had previously practiced as a pediatrician in his home country, potentially limiting the transferability of insights. Future research would benefit from a longitudinal design following medical students, specifically those considering non– hospital-based specialties, as they actively make career choices. This approach would provide real-time insight into their decision-making processes and the factors influencing these trajectories.

## Conclusion

This study shows that career decision-making in medical education is a dynamic, iterative process of testing and negotiating PE fit over time. Experiences of misfit and fit—particularly during clerkships—not only shape specialty preferences but may also prompt deeper reflection on career intentions, including what it means to be a doctor. Our findings underscore the importance of recognizing multiple legitimate professional trajectories within medicine and of supporting students in actively exploring fit across diverse clinical contexts. By fostering reflective practice and making implicit cultural messages explicit, medical schools can better support students in developing sustainable, personally meaningful career paths, while potentially reducing stress surrounding career choice and aligning career decisions with evolving healthcare needs.

## Supporting information

Interview Guide appendix 1

## Data Availability

All data produced in the present work are contained in the manuscript

## Acknowledgements

We wish to thank all participating trainees for participating in this research and Tjarda van Westerop for helping with the transcribing.

## References

1. Agyeman-Manu K, Ghebreyesus TA, Maait M, Rafila A, Tom L, Lima NT, et al. Prioritising the health and care workforce shortage: protect, invest, together. Lancet Glob Health. 2023;11(8):e1162– e4.10.1016/S2214-109X(23)00224-3

2. Gennissen L S-JK, van Exel J, Fluit L, de Graaf J, de Hoog M. Career orientations of medical students: A Qmethodology study. PLoS One. 2021;16(5).10.1371/journal.pone.0249092

3. Fris DAH, van Vianen AEM, Koen J, de Hoog M, de Pagter APJ. Medical students’ career decision-making stress during clinical clerkships. Perspect Med Educ. 2022;11(6):350– 8.10.1007/s40037-022-00734-8

4. Cleland JA, Johnston PW, Anthony M, Khan N, Scott NW. A survey of factors influencing career preference in new-entrant and exiting medical students from four UK medical schools. BMC Med Educ. 2014;14:151.10.1186/1472-6920-14-151

5. Rachoin JS, Vilceanu MO, Franzblau N, Gordon S, Cerceo E. How often do medical students change career preferences over the course of medical school? BMC Med Educ. 2023;23(1).10.1186/s12909-023-04598-2

6. Pfarrwaller E, Maisonneuve H, Laurent C, Abbiati M, Sommer J, Baroffio A, et al. Dynamics of Students’ Career Choice: a Conceptual Framework-Based Qualitative Analysis Focusing on Primary Care. J Gen Intern Med. 2024(39):1544–55.10.1007/s11606-023-08567-9

7. Kaminski A, Falls G, Parikh PP. Clerkship Experiences During Medical School: Influence on Specialty Decision. Med Sci Educ. 2021;31(3):1109–14.10.1007/s40670-021-01281-3

8. Pfarrwaller E, Voirol L, Piumatti G, Karemera M, Sommer J, Gerbase MW, et al. Students’ intentions to practice primary care are associated with their motives to become doctors: a longitudinal study. BMC Med Educ. 2022;22(1):30.10.1186/s12909-021-03091-y

9. Querido SJ, Vergouw D, Wigersma L, Batenburg RS, De Rond ME, Ten Cate OT. Dynamics of career choice among students in undergraduate medical courses. A BEME systematic review: BEME Guide No. 33. Med Teach. 2016;38(1):18–29.10.3109/0142159x.2015.1074990

10. Gutiérrez-Cirlos C, Naveja JJ, García-Minjares M, Martínez-González A, Sánchez-Mendiola M. Specialty choice determinants among Mexican medical students: a cross-sectional study. BMC Med Educ. 2019;19(1):420.10.1186/s12909-019-1830-5

11. Abbiati M, Nendaz MR, Cerutti B, Brodmann Mäder M, Spinas GA, Vicente Alvarez D, et al. Exploring Medical Career Choice to Better Inform Swiss Physician Workforce Planning: Protocol for a National Cohort Study. JMIR Res Protoc. 2024;13:e53138.10.2196/53138

12. Bland CJ, Meurer LN, Maldonado G. Determinants of primary care specialty choice: a non-statistical meta-analysis of the literature. Acad Med. 1995;70(7):620– 41.10.1097/00001888-199507000-00013

13. Passi V, Johnson S, Peile E, Wright S, Hafferty F, Johnson N. Doctor role modelling in medical education: BEME Guide No. 27. Med Teach. 2013;35(9):e1422– 36.10.3109/0142159x.2013.806982

14. Passi V, Johnson N. The hidden process of positive doctor role modelling. Med Teach. 2016;38(7):700–7.10.3109/0142159x.2015.1087482

15. Grasreiner D, Dahmen U, Settmacher U. Specialty preferences and influencing factors: a repeated cross-sectional survey of first-to sixth-year medical students in Jena, Germany. BMC Med Educ. 2018;18(1):103.10.1186/s12909-018-1200-8

16. Velgan M, Vajer P, Michels NR, Einasto M, Kalda R. Factors influencing medical students career intentions in Flanders, Estonia, and Hungary: a multivariable analysis. BJGP Open. 2025;9(2):BJGPO.2024.0087.10.3399/bjgpo.2024.0087

17. Kristof-Brown A. L. SCK. Goal congruence in project teams: Does the fit between members’ personal matery and performance goals matter? J Appl Psychol. 2001(86):1083– 95.10.1037/0021-9010.86.6.1083

18. Chuang Aichia SC-T, Judge Timothy A. Development of a Multidimensional Instrument of Person–Environment Fit: The Perceived Person–Environment Fit Scale (PPEFS). Appl Psychol. 2016;65(1):66–98.10.1111/apps.12036

19. Andela Marie DvdM. A Comprehensive Assessment of the Person–Environment Fit Dimensions and Their Relationships With Work-Related Outcomes. J Career Dev. 2019(46):567–82

20. Xiao Y, Dong M, Shi C, Zeng W, Shao Z, Xie H, et al. Person–environment fit and medical professionals’ job satisfaction, turnover intention, and professional efficacy: A cross-sectional study in Shanghai. PLoS One. 2021;16(4):e0250693.10.1371/journal.pone.0250693

21. Vleugels W, Verbruggen, M., De Cooman, R., & Billsberry. A systematic review of temporal person-environment fit research: Trends, developments, obstacles, and opportunities for future research. J Organ Behav. 2022;44(2):376–98.10.1002/job.2607

22. Dirksen PP, Cillekens B, Hoogsteder M, Soethout M. Shortage of youth health physicians: what is the solution?: A study of characteristics of the profession of youth health physician in the three northern provinces of the Netherlands. TSG. 2025;103(4):106–14.10.1007/s12508-025-00467-z

23. Pfarrwaller E, Audétat M-C, Sommer J, Maisonneuve H, Bischoff T, Nendaz M, et al. An Expanded Conceptual Framework of Medical Students’ Primary Care Career Choice. Acad Med. 2017;92(11):1536–42.10.1097/acm.0000000000001676

24. Memari Fard D. Amberscript (2025). Transcription Guidelines (2025). Taxonomy, Language-Specific Features, and Positioning in Research & Practice.v2025.11–1.10.5281/zenodo.17700610

25. Varpio L, Paradis E, Uijtdehaage S, Young M. The Distinctions Between Theory, Theoretical Framework, and Conceptual Framework. Acad Med. 2020;95(7):989– 94.10.1097/acm.0000000000003075

26. Thomas A, Kinston, R., Yardley, S., McKinley, R. K., & Lefroy, J. How do medical schools influence their students’ career choices? A realist evaluation. Med Educ Online. 2024;29.10.1080/10872981.2024.2320459

27. Zhao Y, Mbuthia D, Blacklock C, Gathara D, Nicodemo C, Molyneux S, et al. How do foundation year and internship experience shape doctors’ career intentions and decisions? A meta-ethnography. Med Teach. 2023;45(1):97–110.10.1080/0142159x.2022.2106839

28. Pfarrwaller E, Laurent C, Sommer J, Baroffio A, Haller DM, Maisonneuve H. ‘I felt I belonged’: A qualitative study of role modelling and team integration as key drivers of primary care career choice. Eur J Gen Pract. 2025;31(1):2527143.10.1080/13814788.2025.2527143

29. Singh A, Alberti H. Why UK medical students change career preferences: an interview study. Perspect Med Educ. 2021;10(1):41–9.10.1007/s40037-020-00636-7

30. Ladha M, Bharwani A, McLaughlin K, Stelfox HT, Bass A. The effect of white coats and gender on medical students’ perceptions of physicians. BMC Med Educ. 2017;17(1):93.10.1186/s12909-017-0932-1

31. Apperloo-Swiersema K HJ, Duvivier R. Considerations of Medical Students for a Career in Public Health. Ned Tijdschr Geneeskd. 2026;170

32. Larrotta SP RE, Correa DN, Peñuela CJ, Tapia AR. Effects of the Hidden Curriculum in Medical Education: Scoping Review. Jmir Medical Education. 2025;11.10.2196/68481

33. Hafferty FW, Franks R. The hidden curriculum, ethics teaching, and the structure of medical education. Acad Med. 1994;69(11):861–71.10.1097/00001888-199411000-00001

34. Chapa H, Dickey D, Milman R, Hagar C, Kintzer J. A Novel Curricular Design Exposing Clinical Medical Students to the Hidden Curriculum. Med Sci Educ. 2022;32:17– 9.10.1007/s40670-021-01479-5

35. Brown MEL CO, Heybourne A, Finn GM. Exploring the Hidden Curriculum’s Impact on Medical Students: Professionalism, Identity Formation and the Need for Transparency. Med Sci Educ. 2020;24(3):1107–21.10.1007/s40670-020-01021-z

36. Holmes CL, Harris IB, Schwartz AJ, Regehr G. Harnessing the hidden curriculum: a four-step approach to developing and reinforcing reflective competencies in medical clinical clerkship. Adv in Health Science. 2015;20:1355–70.10.1007/s10459-014-9558-9

37. Muyselaar-Jellema JZ, Querido SJ. Twelve tips for having more meaningful conversations with medical students on specialty career choice. Med Teach. 2024;46:617– 20.10.1080/0142159X.2023.2280114

