## Supplementary material for "Where Do I Belong? Searching for fit in an unseen specialty: medical students’ paths to Youth Health Care": Interview Guide appendix 1

**Appendix 1: Interview Guideline**

| **Interview phase** | **Topic** | **Probing questions** |
|---|---|---|
| **Welcome and information** | | |
| Describing career decision process / trajectory | You are now a youth health care trainee; can you describe the process through which you decided to pursue youth health care as a specialty? | - Can you tell me about your experiences working as a physician after medical school?<br>- What do you find appealing or interesting about working in Youth Health Care (YHC)?<br>- Can you give a specific example of a situation in which you made a meaningful difference for a child or family as a youth health physician?<br>- How did you decide to pursue specialty training?<br>- Can you tell me what made you choose for the youth health care specialty? |

| | | |
|---|---|---|
| Influences on career intention / career choice | Can you describe the factors, experiences, or people that have influenced the career choices you've made? | - Can you describe a situation or circumstance? |
| | What influence did medical school have on your decision to pursue your current specialty? | - What was the influence of the clerkships on your specialty choice?<br>- Did you undertake a clerkship in Youth Health Care (YHC)? In what other ways were you introduced to the field during medical school? |
| | Can you tell me about the steps you took to explore and choose the specialty you wanted to pursue? | - Can you give examples of what you did to help make your specialty choice? |
| | In hindsight, what would have helped you in your decision process? | |

| | |
|---|---|
| | Were there certain people that were influential or important to you in making your career choice? |
| **End of the interview** | Summary<br>Is there anything relevant that you want to add to this conversation?<br>Do you have any questions? |
| **Conclusion** | Thank the participant |
